# Cerebrospinal fluid in COVID-19 neurological complications: no cytokine storm or neuroinflammation

**DOI:** 10.1101/2021.01.10.20249014

**Authors:** Maria A. Garcia, Paula V. Barreras, Allie Lewis, Gabriel Pinilla, Lori J. Sokoll, Thomas Kickler, Heba Mostafa, Mario Caturegli, Abhay Moghekar, Kathryn C. Fitzgerald, Hopkins Neuro-COVID-19 Group, Carlos A. Pardo

**Affiliations:** Department of Neurology, Johns Hopkins University School of Medicine, Baltimore, Maryland; Bloomberg School of Public Health, Johns Hopkins University School of Medicine, Baltimore, Maryland; Fundacion Valle de Lili, Cali, Colombia; Department of Pathology, Johns Hopkins University School of Medicine, Baltimore, Maryland

**Author notes:** Corresponding author: Carlos A. Pardo, M.D., Johns Hopkins University School of Medicine 600 North Wolfe Street, 627 Pathology Bld, Baltimore, MD, 21287.

**Keywords:** SARS-CoV2, COVID-19, neuroinflammation, cerebrospinal fluid, cytokines, encephalopathy, cytokine storm

## Abstract

**BACKGROUND:** Neurological complications occur in COVID-19. We aimed to examine cerebrospinal fluid (CSF) of COVID-19 subjects with neurological complications and determine presence of neuroinflammatory changes implicated in pathogenesis.

**METHODS:** Cross-sectional study of CSF neuroinflammatory profiles from 18 COVID-19 subjects with neurological complications categorized by diagnosis (stroke, encephalopathy, headache) and illness severity (critical, severe, moderate, mild). COVID-19 CSF was compared with CSF from healthy, infectious and neuroinflammatory disorders and stroke controls (n=82). Cytokines (IL-6, TNFα, IFNγ, IL-10, IL-12p70, IL-17A), inflammation and coagulation markers (high-sensitivity-C Reactive Protein [hsCRP], ferritin, fibrinogen, D-dimer, Factor VIII) and neurofilament light chain (NF-L), were quantified. SARS-CoV2 RNA and SARS-CoV2 IgG and IgA antibodies in CSF were tested with RT-PCR and ELISA.

**RESULTS:** CSF from COVID-19 subjects showed a paucity of neuroinflammatory changes, absence of pleocytosis or specific increases in pro-inflammatory markers or cytokines (IL-6, ferritin, or D-dimer). Anti-SARS-CoV2 antibodies in CSF of COVID-19 subjects (77%) were observed despite no evidence of SARS-CoV2 viral RNA. A similar increase of pro-inflammatory cytokines (IL-6, TNFα, IL-12p70) and IL-10 in CSF of COVID-19 and non-COVID-19 stroke subjects was observed compared to controls. CSF-NF-L was elevated in subjects with stroke and critical COVID-19. CSF-hsCRP was present almost exclusively in COVID-19 cases.

**CONCLUSION:** The paucity of neuroinflammatory changes in CSF of COVID-19 subjects and lack of SARS-CoV2 RNA do not support the presumed neurovirulence of SARS-CoV2 or neuroinflammation in pathogenesis of neurological complications in COVID-19. Elevated CSF-NF-L indicates neuroaxonal injury in COVID-19 cases. The role of CSF SARS-CoV2 IgG antibodies is still undetermined.

**FUNDING:** This work was supported by NIH R01-NS110122 and The Bart McLean Fund for Neuroimmunology Research.

## INTRODUCTION

Central and peripheral nervous system disorders can develop in patients with severe acute respiratory syndrome coronavirus 2 (SARS-CoV2) infection, during acute and/or postinfectious phases of coronavirus disease 2019 (COVID-19).(1, 2) These disorders are influenced by patient age, sex and pre-existing comorbidities and are mainly represented by cerebrovascular pathologies and encephalopathies (3-7). The so-called “COVID-19 encephalitis”(8), acute disseminated encephalomyelitis (ADEM)(9), cranial neuropathies(10) and Guillain-Barré syndrome(11, 12) have also been described as well as a high frequency of headache(13-15). There are several unanswered questions regarding the pathogenesis of these neurological complications including concerns about the neuro-invasiveness or neurovirulence of SARS-CoV2, the role of neuroinflammation and the effects of the “cytokine storm” on the central nervous system (CNS). Studies focused on the analysis of cerebrospinal fluid (CSF) in COVID-19 infection have outlined a diversity of CSF findings that lack specific profiles associated with the neurological symptoms(16-21). Interestingly, IgG antibodies against SARS-CoV2 spike protein have been found in the CSF of eight patients with encephalopathy(22), and other case reports have described changes suggestive of an inflammatory process(23, 24) and neuronal damage(25, 26). In the present study, we explore the possible pathogenesis of COVID-19 associated neurological complications by investigating for markers of neuroinflammation, the presence of SARS-CoV2 RNA and antibodies to SARS-CoV2. We additionally compared the CSF from subjects with COVID-19 associated neurological complications with CSF of non-COVID-19 patients with stroke, neurological infections and neuroinflammatory disorders, to compare the immune profiles which may allow the recognition of common pathogenic pathways.

## RESULTS

### Patient clinical characteristics

A cross-sectional study to investigate immune and neuroinflammatory changes which may be associated with pathogenesis of neurological involvement was performed in the CSF of 18 adult COVID-19 patients with neurological manifestations and compared to those of 14 age matched healthy and 68 non-COVID-19 neurological disease controls. Neurological manifestations in COVID-19 were categorized in three diagnostic groups: stroke (N=7), encephalopathies (N=6) and headaches/others (N=5). The clinical and neuroimaging features, and systemic inflammatory markers for all COVID-19 subjects are described in Table 1. The temporal profile of infection and neurological symptoms, the time of diagnosis by nucleic acid amplification test (NAAT) by RT-PCR for SARS-CoV2 in nasopharyngeal swab (NS) [NS-NAAT], CSF sampling and clinical events such as intubation/extubation are outlined in Figure 1. The median age of the patients was 56 years (IQR 32-69). Ten patients were male (56%). Eight (44%) patients were classified with critical illness, 5 (28%) with severe illness, 4 (22%) with moderate illness and 1 with mild illness. Six patients (33%) were diagnosed with COVID-19 hyperinflammatory syndrome (C-HIS)(27) of whom four had encephalopathy. The diagnosis of COVID-19 was established by NS-NAAT in 16 patients, and two patients were diagnosed based on positive serum anti-SARS-CoV2 IgG and IgA antibodies. The NS-NAAT CT value, a presumptive indicator of the magnitude of viral infection(28, 29), was established in 12 patients, 7 of them with CT <30. Four (22%) of the 18 patients died, while 13 (72%) improved. Of the 18 patients, 7 (39%) were diagnosed with stroke, 6 (33%) had encephalopathy and 5 (27%) were diagnosed as headache/ other. Six of 18 (33%) patients had 3 or more comorbidities while 9 (66%) had at least one comorbidity. Overall, the median time from onset of COVID-19 to neurological symptoms was 0.5 days (IQR 0-10.5). Interestingly, in nine patients (50%), neurological manifestations were part of the initial clinical presentation of COVID-19 symptoms (3 stroke, 4 encephalopathy and 2 headache). CSF control samples were classified as: 1) healthy controls (n=14); 2) acute meningitis (n=12), 3) acute viral encephalitis (e.g., herpes simplex, varicella-zoster encephalitis)(30) (n=11), 4) autoimmune encephalitis(31) (n=14), 5) neuromyelitis optica (NMO)(32) (n=11), 6) neurosarcoidosis (33) (n=12), and 7) stroke (n=8), which included subjects with ischemic stroke (34) preceding the COVID-19 period. Demographic characteristics for the control groups are described in supplemental Table 1.

**Table 1.**
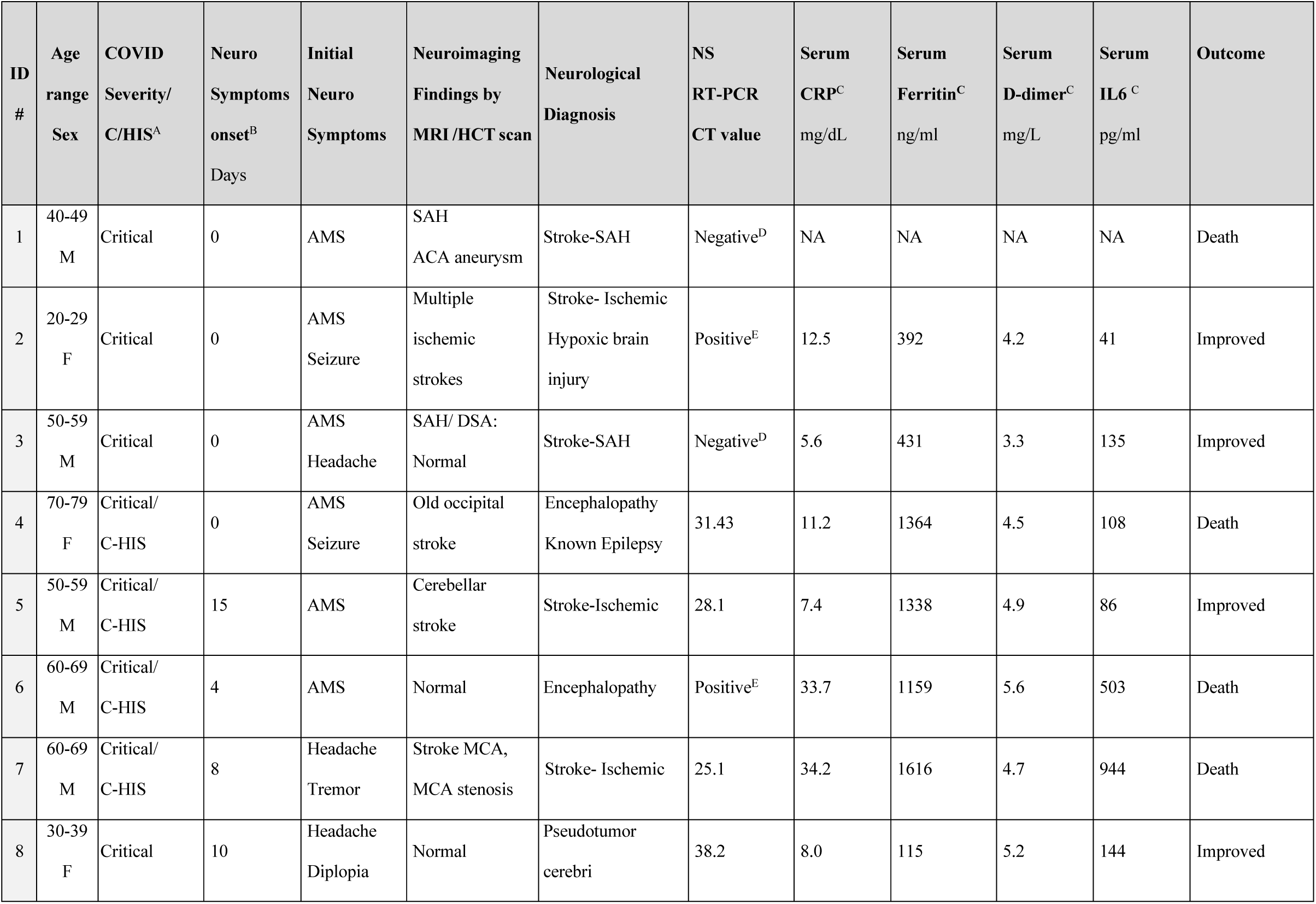
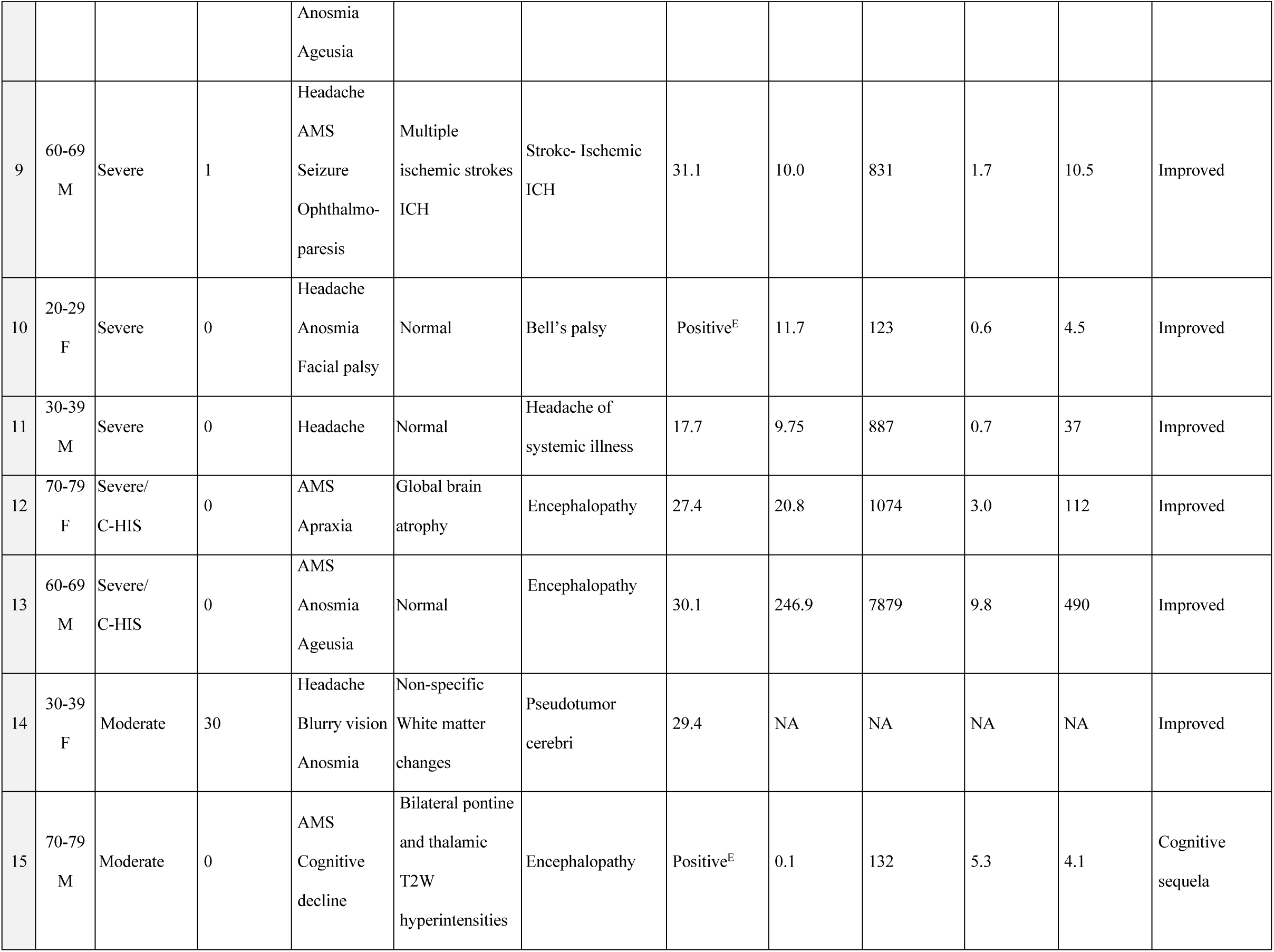
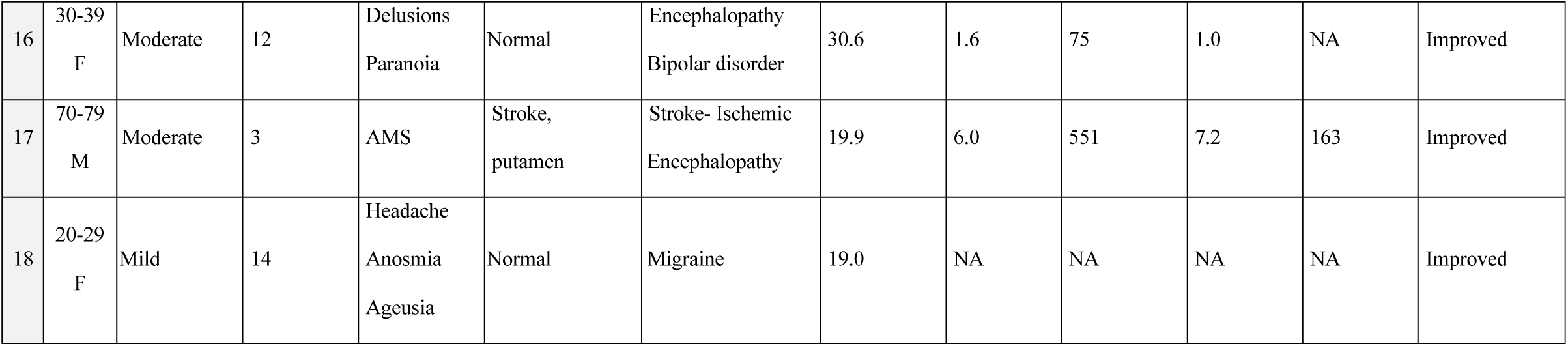
Clinical characteristics of subjects with COVID-19 neurological manifestations.

**Figure 1.**
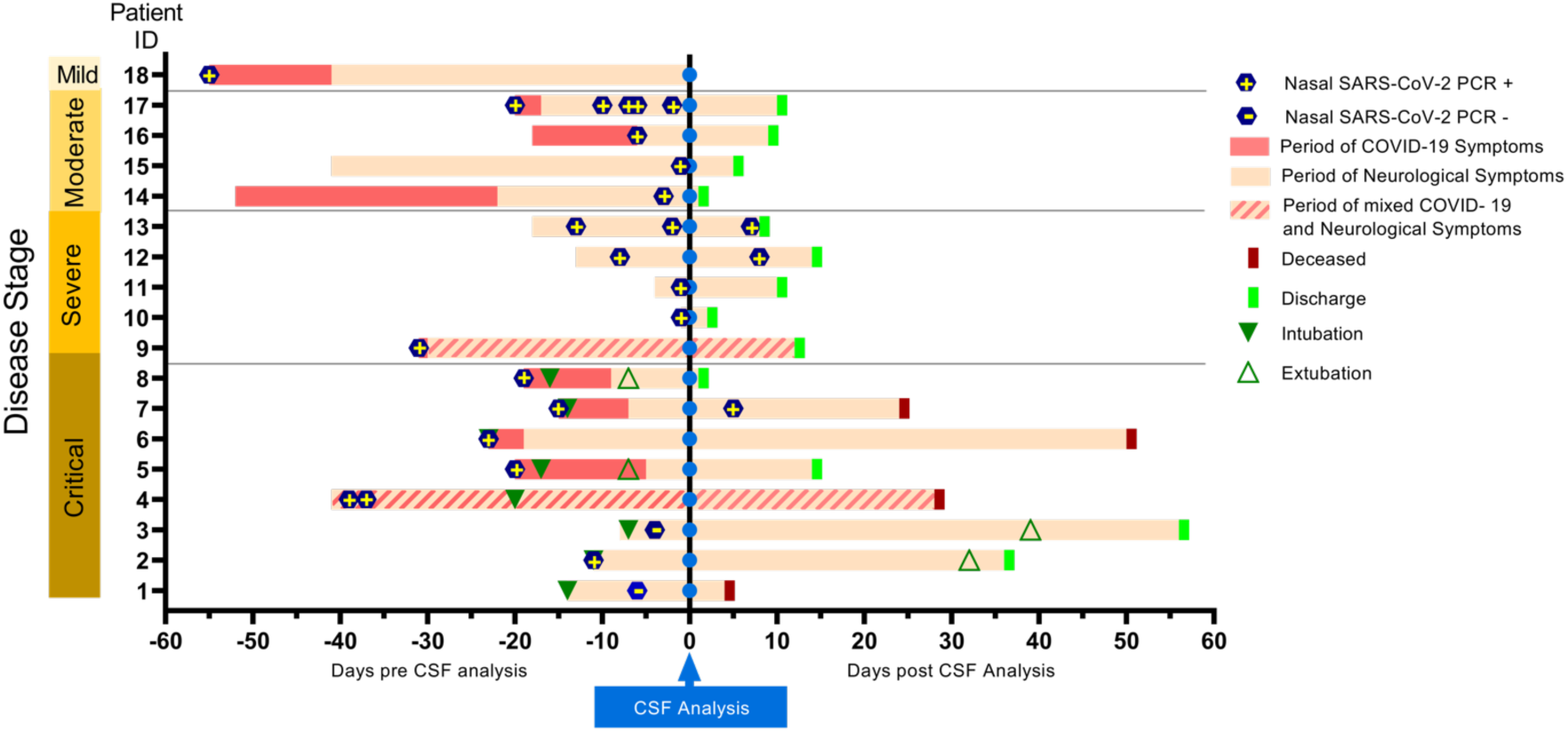
Timeline of clinical features in patients with COVID-19 with neurological complications. Temporal profile of COVID-19 and neurological symptoms as related with the time of CSF analysis (vertical blue line) for the 18 subjects included in the study. Patients were classified based on the NIH disease severity classification (56). Eight subjects presented with neurological symptoms as the first manifestation of COVID-19 (light pink bar), eight exhibited systemic illness symptoms preceding neurological symptoms (dark pink bar) and two presented with mixed neurological and systemic symptoms (diagonal stripes).

### CSF characteristics

CSF features for the COVID-19 subjects are summarized in Table 2. The CSF was collected within 8 days of the first positive NS-NAAT in 8 patients (44%, median 4 (IQR 1 - 6) while other 10 patients (56%, median 20 (IQR 13 - 27) had a CSF collection 9 days or more after COVID-19 diagnosis. There was no evidence of CSF pleocytosis except in 4 subjects in which the increase of white cell count (WCC) was likely due to cross contamination with blood. In 8 subjects, the CSF protein was elevated, four of them likely influenced by an excessive number of red blood cells and/or stroke. The CSF IgG index and CSF/albumin ratio (Qalb) were within normal range in 7 patients where these indexes were tested. No evidence of oligoclonal bands (OCBs) in CSF or corresponding serum was found in 5 subjects where this test was obtained.

**Table 2.**
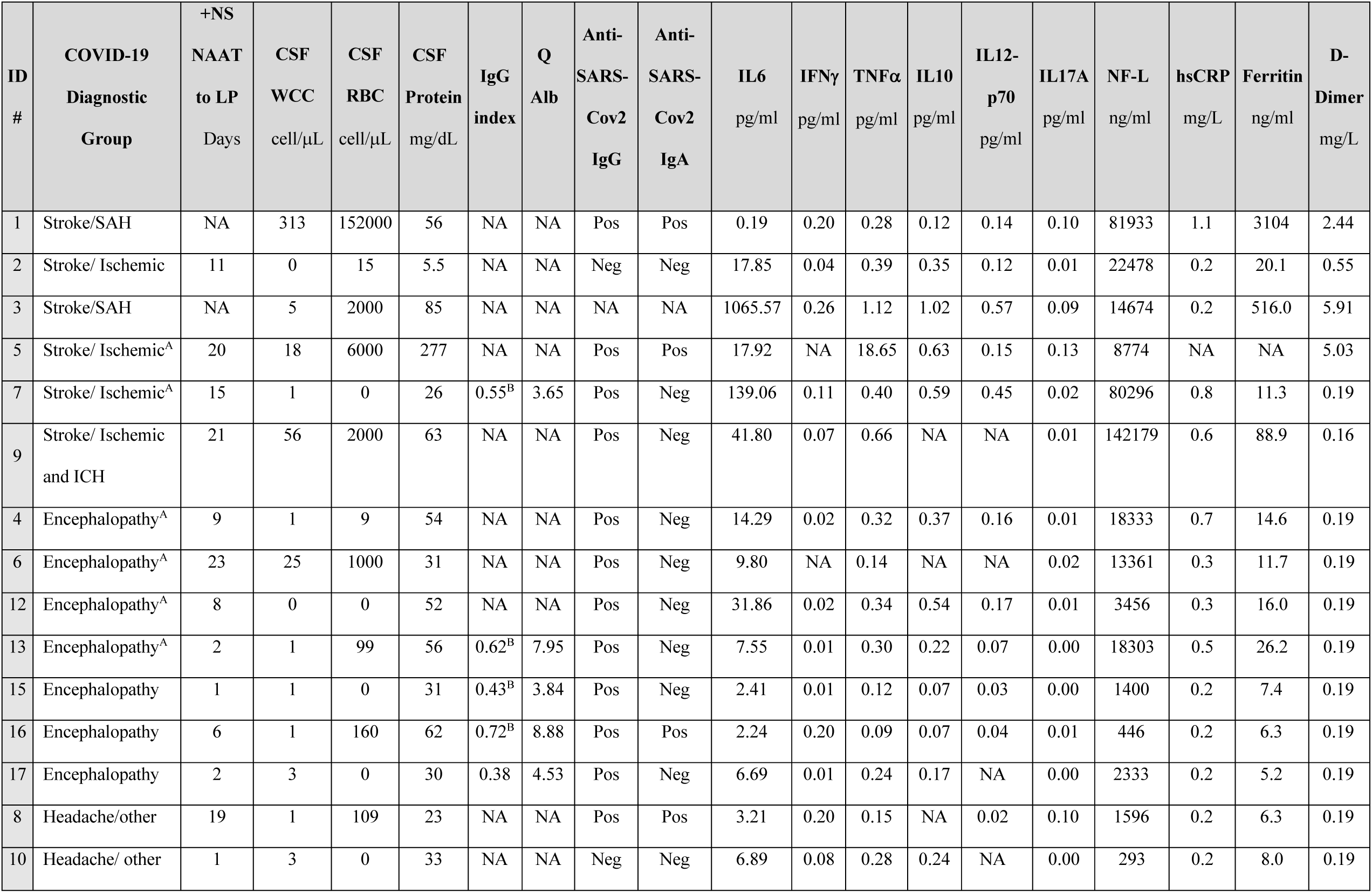
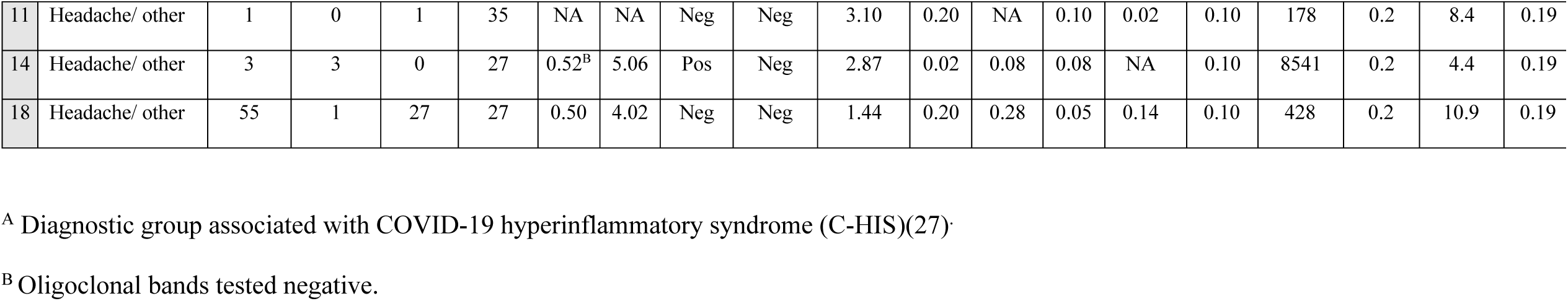
Cerebrospinal fluid neuroinflammatory markers in COVID-19 subjects.

### SARS-CoV2 testing in CSF

We investigated the presence of SARS-CoV2 in all CSF samples from the COVID-19 group. None of the 18 CSF samples from COVID-19 was positive for SARS-CoV2 RNA by RT-PCR. A subset of 7 CSF samples were also tested with Reverse Transcription Droplet Digital PCR (ddPCR) with negative results. We also determined the presence of anti-SARS-CoV2 IgG and IgA antibodies against subunit 1 of the trimeric SARS-CoV2 spike protein. IgG antibodies to SARS-CoV2 spike protein were detected in 13 of 17 (77%) CSF tested while the anti-SARS-CoV2 IgA antibody was detected in 4 of those CSF samples (Table 2, supplemental Figure 1). The titer of the IgG antibody did not correlate with the period between the onset of COVID-19 symptoms and CSF sampling (p=0.53) or period between NS-NAAT diagnosis and CSF sampling (p=0.45). The presence of IgG antibodies to SARS-CoV2 in CSF was observed in all COVID-19 diagnostic groups or disease severity categories. None of the 20 control samples tested for COVID-19 antibodies were positive. In 6 COVID-19 subjects with IgG antibodies, there was almost complete absence of WCC or RBC, although in 7 subjects RBC>50 was noted in CSF as result of either stroke (n=3) or possible traumatic lumbar puncture (n=4)(supplemental Table 2).

### Laboratory findings in CSF of COVID-19 and control groups

To determine the role that immune and pro-inflammatory factors may play in the pathogenesis of COVID-19 neurological complications, we quantified in the CSF the concentrations of cytokines (IL-6, TNFα, IFNγ, IL-10, IL-12p70, IL-17A), inflammation and coagulation markers (high-sensitivity-C Reactive Protein (hsCRP), ferritin, fibrinogen, D-dimer, Factor VIII) and neurofilament light chain (NF-L). We compared the CSF from subjects with COVID-19 associated neurological complications with CSF of non-COVID-19 subjects with neurological infections, neuroinflammatory disorders and stroke to compare the immune profiles which may allow the recognition of common pathogenic pathways. The description of CSF analytes in the COVID-19 diagnostic groups and control groups are included in Table 3. Comparative analysis and statistical outcomes for all CSF analytes is shown in Figure 2. A representative heat map of the P value significance for all analyte comparisons between the COVID-19 diagnostic categories, disease severity and timing of CSF collection with the control groups are described in Figure 3. No significant differences in the CSF WCC and protein concentrations between the three diagnostic categories of the COVID-19 neurological problems were found. As compared with healthy controls, there were no significant differences in the WCC and protein concentration with exception of the COVID-19 headache group which had a significantly lower protein concentration. Overall, the WCC and protein concentrations were lower in COVID-19 cases as compared with neuroinflammatory controls. No significant differences were seen between COVID-19 and non-COVID-19 stroke cases.

**Table 3.** **Cerebrospinal fluid quantification of neuroinflammatory biomarkers in COVID-19 and control groups**

| Analytes<br>Median<br>(IQR) | COVID-19 |  |  |  | Comparison Control Groups |  |  |  |  |  |  |
| --- | --- | --- | --- | --- | --- | --- | --- | --- | --- | --- | --- |
|  | All<br>N=18 | Stroke<br>N=7 | Encephalopathy<br>N=6 | Headache<br>N=5 | Healthy<br>controls<br>N=14 | Acute<br>meningitis<br>N=12 | Acute<br>encephalitis<br>N=11 | Autoimmune<br>encephalitis<br>N=14 | NMO<br>N=11 | Neuro-Sarcoidosis<br>N=12 | Stroke<br>N=8 |
| **WCC**<br>cell/μL | 2<br>(1-5) | 5<br>(1-56) | 1 | 2<br>(1-4) | 1<br>(0-2) | 65<br>(20-203) | 7<br>(1-21) | 3<br>(2-7) | 9<br>(2-74) | 26<br>(2-32) | 3<br>(1-27) |
| **Protein**<br>mg/dL | 34<br>(29.2-56.8) | 56.4<br>(26.2-85.3) | 53<br>(31.3-56.8) | 29.2<br>(27.5-33) | 41.1<br>(33-2-57) | 80.3<br>(49.5-108.5) | 50.8<br>(34-73) | 34.5<br>(21-43) | 38<br>(27-63) | 76.5<br>(43.8-137.8) | 50.5<br>(30.2-73.2) |
| **NF-L**<br>pg/mL | 8657<br>(1400-18333) | 22477<br>(8773-81933) | 8408<br>(1400-18302) | 428<br>(292-1595) | 1105<br>(750-2291) | 6860<br>(2334-16425) | 20560<br>(11243-89277) | 20914<br>(2594-29769) | 8812<br>(1867-11979) | 2697<br>(1168-9380) | 4330<br>(898-35388) |
| **IL-6**<br>pg/mL | 7.22<br>(2.8-17.92) | 17.92<br>(6.69-139.06) | 8.67<br>(2.41-14.29) | 3.09<br>(2.86-3.20) | 3.31<br>(2.32-4.67) | 44.57<br>(3.38-165.17) | 6.51<br>(2.75-28.33) | 3.26<br>(2.62-23.52) | 4.05<br>(2.8-61.93) | 22.12<br>(2.35-188.8) | 37.13<br>(6.45-98.51) |
| **TNFα**<br>pg/mL | 0.27<br>(0.14-0.39) | 0.4<br>(0.28-1.20) | 0.22<br>(0.12-0.32) | 0.21<br>(0.11-0.28) | 0.13<br>(0.11-0.20) | 1.47<br>(0.77-3.11) | 0.72<br>(0.46-0.82) | 0.38<br>(0.17-0.85) | 0.27<br>(0.13-0.30) | 1.68<br>(0.21-7.23) | 0.52<br>(0.27-0.75) |
| **IFNγ**<br>pg/mL | 0.08<br>(0.01-0.20) | 0.09<br>(0.04-0.20) | 0.02<br>(0.01-0.02) | 0.20<br>(0.08-0.20) | 0.18<br>(0.02-0.20) | 0.73<br>(0.20-4.22) | 0.25<br>(0.08-2.06) | 0.20<br>(0.08-0.20) | 0.18<br>(0.02-0.20) | 3.04<br>(0.05-55.49) | 0.08<br>(0.02-0.20) |
| **IL-10**<br>pg/mL | 0.21<br>(0.07-0.54) | 0.47<br>(0.16-0.62) | 0.21<br>(0.07-0.37) | 0.09<br>(0.06-0.17) | 0.08<br>(0.05-0.16) | 0.58<br>(0.45-1.03) | 0.30<br>(0.07-0.87) | 0.13<br>(0.08-0.45) | 0.19<br>(0.07-0.35) | 0.44<br>(0.08-1.29) | 0.64<br>(0.34-0.78) |
| **IL-12p70**<br>pg/mL | 0.13<br>(0.03-0.15) | 0.15<br>(0.14-0.45) | 0.07<br>(0.03-0.16) | 0.03<br>(0.02-0.14) | 0.03<br>(0.02-0.04) | 0.23<br>(0.13-0.68) | 0.08<br>(0.02-0.44) | 0.08<br>(0.03-0.20) | 0.06<br>(0.04-0.09) | 0.19<br>(0.05-0.59) | 0.24<br>(0.06-0.25) |
| **IL-17A**<br>pg/mL | 0.02<br>(0.01-0.10) | 0.02<br>(0.01-0.10) | 0.01<br>(0.00-0.01) | 0.10<br>(0.10-0.10) | 0.10<br>(0.00-0.10) | 0.16<br>(0.09-1.71) | 0.09<br>(0.02-0.14) | 0.10<br>(0.03-0.10) | 0.08<br>(0.03-0.10) | 0.10<br>(0.05-0.17) | 0.03<br>(0.01-0.07) |
| **Ferritin**<br>ng/mL | 11.3<br>(7.4-20.1) | 54.5<br>(11.3-516) | 13.1<br>(7.4-16) | 8<br>(6.3-8.4) | 8.8<br>(7.8-11) | 9.4<br>(6.9-14) | 10.7<br>(5.9-35) | 8.4<br>(4.3-10.7) | 7.4<br>(6.2-9.6) | 7.9<br>(5.5-14) | 9<br>(4.7-46.7) |
| **D-dimer**<br>mg/L | 0.19 | 0.55<br>(0.19-2.44) | 0.19 | 0.19 | 0.19<br>(0.19-0.37) | 0.24<br>(0.19-1.4) | 0.3<br>(0.19-1.14) | 0.19 | 0.19<br>(0.19-0.65) | 1.2<br>(0.33-4.01) | 0.32<br>(0.19-1.44) |
| **D-dimer\***<br>>0.19 mg/L | 4 | 4 | 0 | 0 | 3 | 5 | 3 | 1 | 2 | 7 | 4 |
| **hsCRP\***<br>>0.2 mg/L | 7 | 3 | 4 | 0 | 0 | 0 | 0 | 1 | 0 | 0 | 0 |

**Figure 2.**
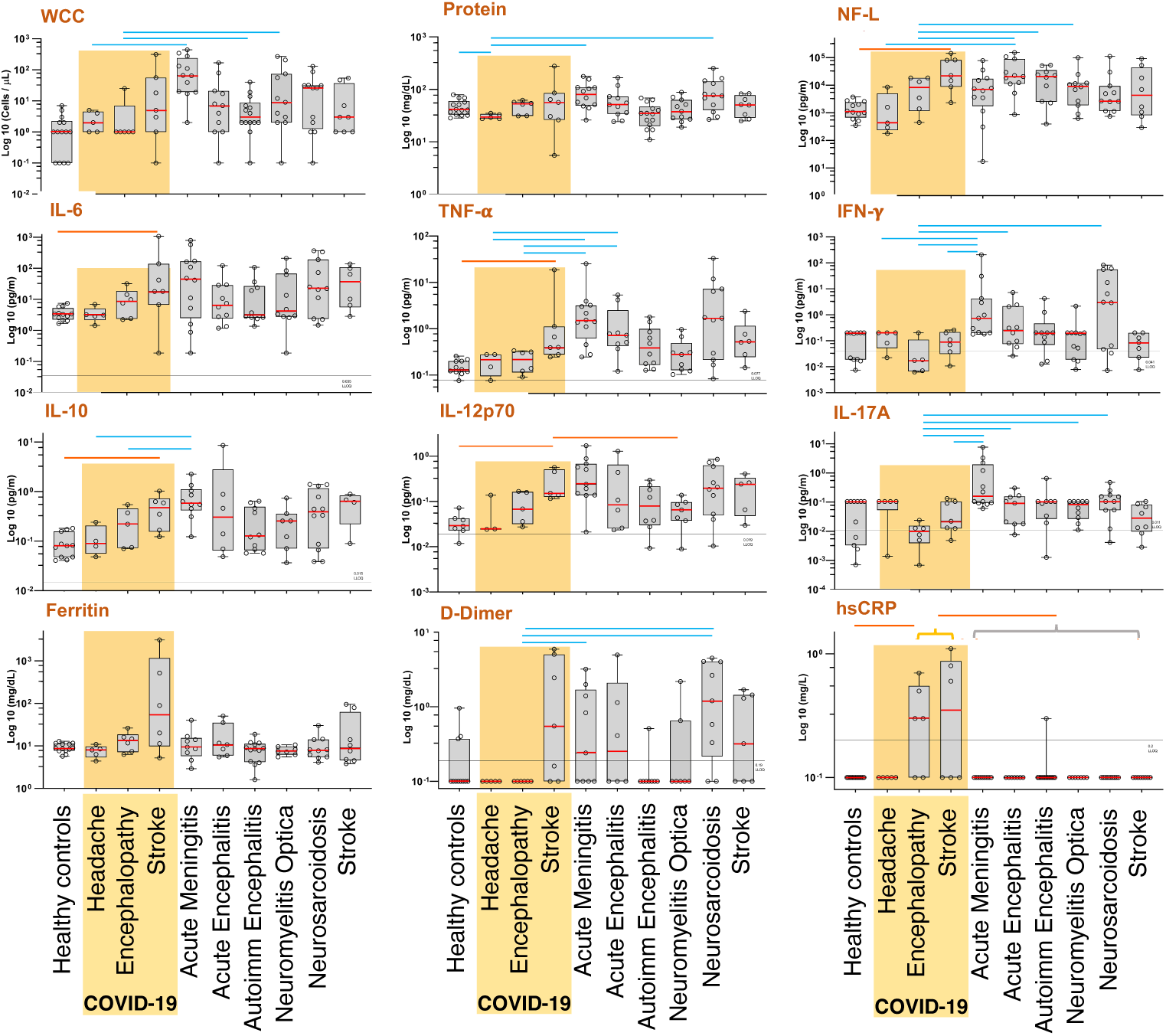
Profile of CSF inflammatory markers in COVID-19 diagnostic groups and controls. Profiles of inflammatory markers in the CSF from 18 COVID-19 subjects categorized by diagnosis (yellow box) as compared with healthy controls (n= 14), acute meningitis (n=12) and encephalitis (n=14), autoimmune encephalitis (n=12), neuromyelitis optica (n=11) and neurosarcoidosis (n=14) and non-COVID-19 strokes (n=8). Boxes indicate the interquartile range and whiskers show the minimum and maximum values for each group, and median (red line in box plot). P> 0.05 is denoted as an orange line when COVID-19 diagnostic group was significantly higher than the control and as a blue line when the control group was significantly higher than the COVID-19 diagnostic category. Significance for D-dimer and hsCRP was obtained by categorical analysis, present or absent. The significance for hsCRP was represented by the COVID-19 stroke and encephalopathy groups (yellow bracket) vs. the healthy controls and the overall disease control groups (gray bracket).

**Figure 3.**
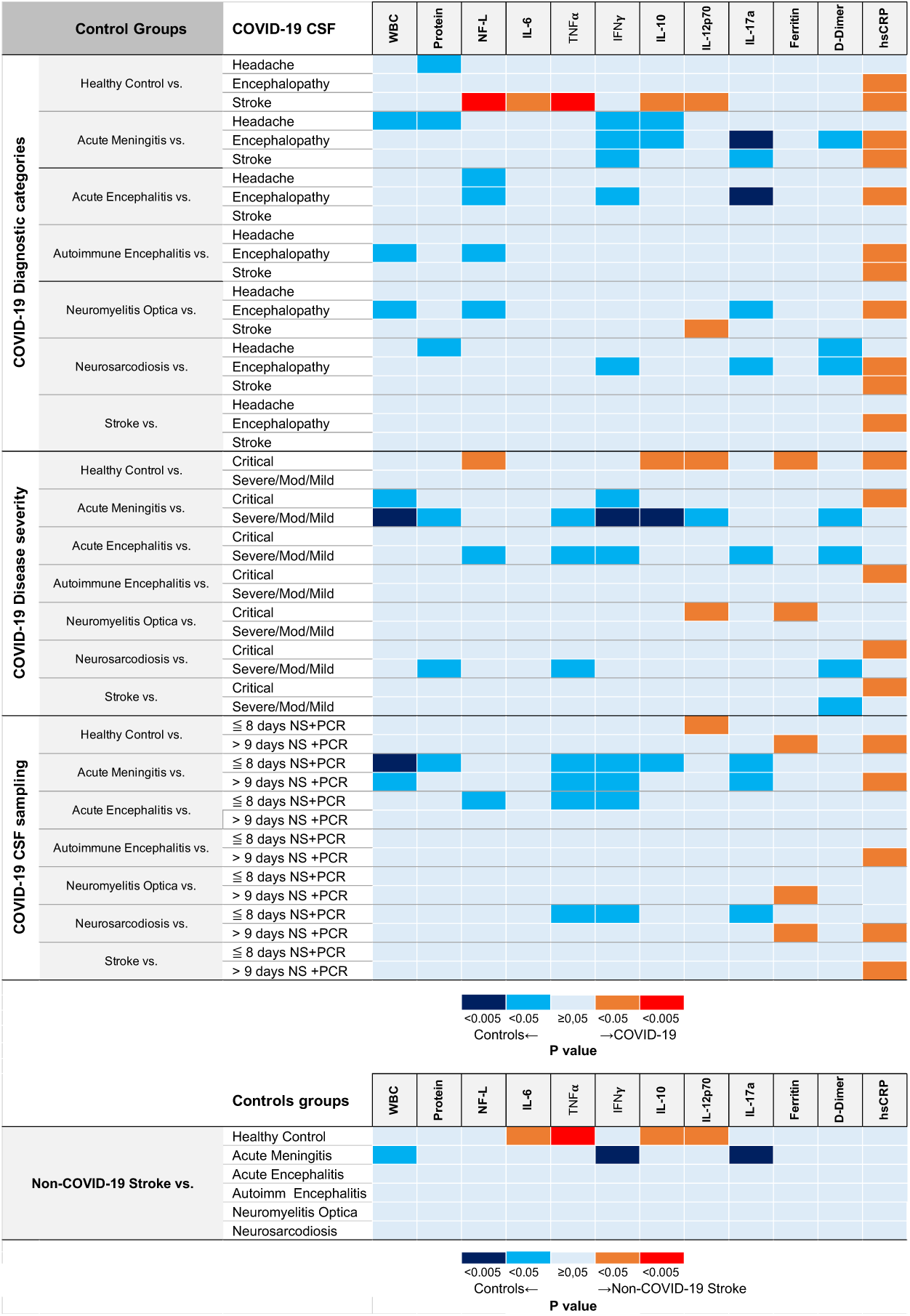
Heatmap of significance analysis of COVID-19 cases vs. controls. Heatmap description of the significance (P<0.05) of the comparative analysis based on (A) COVID-19 CSF diagnostic groups, (B) COVID-19 disease severity, and (C) timing of the CSF sampling in COVID-19 vs. control groups. (D) Heatmap description of comparison of the non-COVID-19 stroke group with other control groups. P< 0.05 is denoted as orange or red line when COVID-19 diagnostic group was significantly higher as control or light and dark blue when the control group was significantly higher than the COVID-19 diagnostic category (A, B and C) or when the non-COVID-19 stroke group was significantly higher (red or orange) or significantly lower (blues) than other controls. Significance for D-dimer and hsCRP was obtained by categorical analysis, present or absent.

### Markers of neuroaxonal degeneration

Quantification of neurofilament light chain (NF-L) in CSF was used as indicator of neuroaxonal damage in COVID-19 and control subjects. When comparing based on severity, CSF NF-L levels were significantly elevated in the critical COVID-19 group compared to the other severity categories. CSF concentrations of NF-L were also markedly elevated in the COVID-19 stroke group as compared with healthy controls (P=< 0.001) and the COVID-19 headache group (P=< 0.01). Although the COVID-19 encephalopathy group had an elevated median NF-L concentration of 8408 pg/mL, compared to 1105 pg/mL of the healthy control group, this difference was not statistically significant. As expected, NF-L concentrations were significantly elevated in the acute encephalitis, autoimmune encephalitis and NMO controls groups as compared with COVID-19 encephalopathy. However, the concentrations of NF-L were equivalent in the COVID-19 stroke group and the control stroke group.

### Cytokine profiles

To establish the role of cytokines in pathogenesis of COVID-19 neurological complications, we determined the CSF concentrations of selected cytokines IL6, TNFα, IFNγ, IL-10, IL12p70 and IL17A described to be involved in severe and critical COVID-19.(35-38) The concentrations, comparative analysis and statistical significance between COVID-19 groups and controls for all 6 cytokines analyzed are described in Table 3, and Figures 2 and 3. The sensitivity of the assay for detection of the of the six cytokines across all CSF diagnostic groups is described in supplemental Table 3. Analysis of cytokines levels within the COVID-19 diagnostic categories showed that only TNFα in the COVID-19 stroke group was significantly increased compared to the headache group (P=0.03). When the COVID-19 diagnostic groups were compared with controls, the COVID-19 stroke group had significant elevated concentrations of IL-6, TNFα, IL-10 and IL-12p70 as compared with the healthy control group. None of the COVID-19 diagnostic groups, including the stroke group, showed any significant increase of cytokines as compared with neuroinflammatory or non-COVID-19 stroke control groups. Instead, significant increased concentrations of selected cytokines such as TNFα and IFNγ were noted in neuroinflammatory groups such as acute meningitis and encephalitis as compared with the COVID-19 stroke and encephalopathy groups. IFNγ and IL-17A concentrations were reduced in COVID-19 CSF as compared with inflammatory control groups such as acute meningitis, encephalitis and neurosarcoidosis (Figure 3A). To determine whether such pattern of cytokines was specific to the COVID-19 stroke group, we analyzed separately the cytokine profiles of the non-COVID-19 stroke group with other control groups. We found the non-COVID-19 stroke group had significant increases in the concentrations of IL6, TNFα, IL-10 and IL-12p70 as compared with the healthy control group, and reduced levels of IFNγ and IL-17A as compared with acute meningitis groups (Figure 3D), a profile similar to the one observed in the COVID-19 stroke group. Cytokines profiles as related with COVID-19 disease severity and timing of CSF collection are summarized in the statistical significance heatmap illustrated in Figure 3. When COVID-19 subjects were categorized by disease severity, the critical illness group (n=8) had significantly increased levels of IL-10 and IL-12p70 when compared with healthy controls (Figure 3B). The timing of CSF collection did not show any significant change in the profile of COVID-19 cytokines with exception of an increased IL-12p70 in the “early” COVID-19 CSF sampling group as compared with healthy controls (Figure 3C). Interestingly, analysis of the COVID-19 subjects with C-HIS (n=6), which is characterized by marked systemic inflammatory response or “cytokine storm”(27), showed that CSF cytokine levels did not have any significant differences with the healthy control group. However, an analysis within the COVID-19 group showed levels of IL-6 and IL-10 were significantly elevated (P=0.02 and 0.01 respectively) while concentrations of INFγ and IL-12p70 were significantly lower (P=0.04 for both cytokines) in the COVID-19 C-HIS cases (n=6) when compared with other non-H-CIS COVID-19 cases (n=12). Interestingly, the concentration of IL-6 in the CSF of COVID-19 cases did not correlate with the corresponding serum IL-6 (P=0.27). The effect of specific treatments (e.g., steroids, antivirals) on CSF inflammatory markers was also evaluated. Of the 18 patients included in our cohort of COVID-19 subjects, only 2 subjects (1 and 2, Table 1) had received steroids during the 5 days preceding the lumbar puncture. None of the COVID-19 patients received any of the experimental drugs used during the period prior to CSF collection.

### Acute phase reactants and coagulation markers

Acute phase reactants such as ferritin, C-reactive protein (CRP) and coagulation markers including D-dimer, fibrinogen and factor VIII, have been associated to be markers of severe and critical COVID-19. (39-42) To investigate if they are associated with mechanisms of neurological involvement in COVID-19, we quantified such markers in the CSF of COVID-19 subjects and controls. Using a high-sensitive CRP (hsCRP) assay, we found that CRP was present almost exclusively in the CSF of COVID-19 subjects as it was detected in 7/18 subjects, 4 COVID-19 encephalopathy and 3 COVID-19 stroke, while only one CSF (from an autoimmune encephalitis subject) of the 82 CSF controls had detectable hsCRP (P= 0.001). CSF hsCRP levels strongly correlated with CRP serum levels (P=0.001, Spearman’s ρ 0.765). CSF hsCRP was present only in critical or severe COVID-19 subjects and was elevated in 5 of 6 subjects with COVID-19 C-HIS. In contrast, while the CSF ferritin had a 100% detection rate in the CSF of all COVID-19 and comparison groups (Table 3), there was not significant difference between the concentrations of CSF ferritin when COVID-19 diagnostic groups were compared with healthy, neuroinflammatory or stroke controls. However, an analysis within the COVID-19 diagnostic categories showed the stroke group had significantly elevated levels of CSF ferritin as compared with the headache group (P=0.04). Interestingly, analysis of COVID-19 categorized by disease severity showed that CSF ferritin levels in severe COVID-19 subjects were significantly increased as compared with healthy controls and NMO cases (Figure 3). Similar findings were observed in the “late” CSF collection group. However, those observations were likely biased by the inclusion of cases of subarachnoid hemorrhage and ICH (e.g., cases 1,3 and 9), clinical situations well known for an increased CSF ferritin(43). Importantly, CSF ferritin levels did not parallel serum ferritin levels (*r* 0.206, P=0.46), one of the most important markers of systemic immune activation in COVID-19.(42, 44) Among markers of coagulation, CSF D-dimer was present in 4 COVID-19 stroke subjects and in 25 of the CSF controls (P=0.06). CSF D-dimer was not significantly different between COVID-19 groups and healthy controls. Instead, CSF D-dimer in cases of acute meningitis and neurosarcoidosis was significantly increased as compared with COVID-19 encephalopathy cases. Markers of coagulation such as fibrinogen and Factor VIII were undetectable in the CSF of COVID-19 subjects and comparison controls.

## DISCUSSION

In contrast with prevailing assumptions, our study reveals a paucity of neuroinflammatory changes as reflected by the lack of specific increases in CSF pro-inflammatory cytokines or markers of systemic inflammation such as IL-6, ferritin, or D-dimer as typically seen in serum of COVID-19 patients. These findings paralleled a lack of cellular responses like pleocytosis or other markers of immunological activity within the CNS such as increase in IgG index or OCBs. The absence of meaningful neuroinflammatory changes in the CSF of COVID-19 cases is further demonstrated when it is compared to CSF from acute infectious or autoimmune neuroinflammatory pathologies that show significantly greater inflammatory changes. Our study also reveals: 1) a prominent presence of SARS-CoV2 antibodies in absence of SARS-CoV2 viral RNA in the CSF of subjects with COVID-19 neurological complications; 2) elevated levels of CSF hsCRP in a subset of subjects with critical and severe COVID-19 illness; 3) marked increase in CSF NF-L, a marker of neuroaxonal injury in cases of critical COVID-19 illness and stroke; 4) an essentially equivalent increase of a subset of pro-inflammatory cytokines in the CSF in cases of COVID-19 stroke and non-COVID-19 stroke subjects.

The lack of CSF pleocytosis in COVID-19 subjects, normal protein, and absence of abnormalities in IgG index or Q(Alb), concurs with other studies (16-20). This paucity of CSF inflammatory changes undermines the hypothesis that conventional neuroinflammatory or encephalitic processes play roles in the pathogenesis of the most common neurological complications associated with COVID-19 that were studied here. Furthermore, our case-control study approach of CSF immune markers showed that in subjects with COVID-19 who experienced complications such as stroke or encephalopathy, most of the CSF changes appear to be determined by other pathologies such as ischemic disease likely driven by systemic or vascular factors that influence the development of such brain pathologies rather than primary neuroimmune mediated processes. An important caveat is that we did not test the full spectrum of reported neurologic complications in COVID-19, including multiple cranial neuropathies, ADEM, or GBS, and CSF analysis in those conditions may show different results.

We failed to detect SARS-CoV2 viral RNA in the CSF of all COVID-19 subjects examined, concurring with other studies(16, 18-21). The lack of SARS-CoV2 RNA in the CSF may be interpreted as lack of neuro-invasiveness, absence of active viral replication or simply a relatively low viral trafficking into the CNS. Although the detection of RNA viruses in CSF has been historically challenging in some viral disorders of the CNS,(45) the absence of viral RNA along with the lack of pleocytosis and other inflammatory changes in the CSF of COVID-19 patients supports the conclusion that there is not an active trafficking of SARS-CoV2 into the CNS causing neuroinflammation. This distinguishes it from other RNA viruses like poliomyelitis, enterovirus, West-Nile virus that are difficult to detect but produce blatant signs of neuroinflammation in the CSF(45-47).

A noteworthy observation in our study is a high prevalence (77%) of SARS-CoV2 spike IgG antibodies in the CSF of COVID-19 cases. Given the absence of viral RNA in the CSF, the lack of pleocytosis which may facilitate B-cells trafficking into the CNS, and absence of intrathecal IgG production (e.g., IgG index, OCBs), CSF antibodies to SARS-CoV2 likely originate from serum and then transfer into the CNS despite an otherwise intact blood-CSF barrier, as occurs in other CNS pathologies.(48) Alternatively, stroke or ischemic changes may have altered the CSF-blood brain barrier to facilitate permeability of IgG antibodies. Presence of SARS-CoV2 antibodies in CSF has been also reported by previous studies which raises the possibility that they are directly pathogenic in the neurological complications of COVID-19(22, 49). In a prior study of eight encephalopathic or comatose patients with COVID-19, a high titer of SARS-CoV2 IgG was observed in 50% of them, with one patient showing an increased IgG index(22).

Another study of 3 patients with COVID-19 critical illness found evidence of SARS-CoV2 IgM in the CSF(49). In our study, the presence of SARS-CoV2 antibodies was not associated with specific COVID-19 diagnostic group or disease severity. Although 7 of 13 COVID-19 subjects with positive SARS-CoV2 antibodies in the CSF had potential “contamination” from blood derived from SAH, stroke or a traumatic lumbar puncture, six - did not exhibit any meaningful WCC or RBC presence. Future studies looking for sites of SARS-CoV2 antibody cross reactivity in the CNS, potential neurological effects or pathologic potential in animal models, would be helpful to clarify this question.

Notably, our study showed an impressive lack of pro-inflammatory cytokines in the CSF of subjects with COVID-19 neurological problems. With the exception of COVID-19 stroke cases, COVID-19 encephalopathy or headache cases did not show a noticeable pro-inflammatory cytokine response in the CSF as compared with controls. This observation suggests that local increases of pro-inflammatory cytokines are unlikely the pathogenic factors associated with the neurological symptoms observed in COVID-19 encephalopathy or headache. Only the CSF of the COVID-19 stroke group appeared to have significant increase in IL-6, TNFα, IL10 and IL-12p70 as compared with the healthy control group and the COVID-19 headache group. However, these cytokine increases were largely equivalent in the non-COVID-19 stroke controls, suggesting that the cytokine increases in the brain of COVID-19 stroke subjects are likely driven by stroke and ischemic pathology (50-52) rather than specific neuroinflammatory changes associated with COVID-19. Furthermore, CSF from subjects with C-HIS showed not increases in proinflammatory cytokines as compared with healthy controls although increases in IL-6 and IL-10 and lower IFNγ and IL12p70 differentiate them from non-C-HIS subjects. NF-L, a marker of neuroaxonal injury, was increased in stroke and critical COVID-19 cases, and within the COVID-19 cases, it was elevated in stroke and encephalopathy compared to the headache group. Such observation emphasizes the fact that in critical and severe COVID-19 encephalopathy cases, a process of neuronal injury occurs even in absence of neuroimaging evidence of stroke or ischemic injury and may suggest that such neuronal damage is associated with ischemia, hypoxia and systemic illness. Such finding concurs with previous observations of elevation of neuronal and glial proteins in the CSF in the setting of critical COVID-19 illness and support the approach for using such proteins as potential biomarkers of disease process and outcome in future serological studies. (53, 54)

Remarkably, our study demonstrated absent parallel increase in CSF of markers such as IL-6, ferritin, D-dimer or coagulation factors as it has been observed in the serum of COVID-19 patients. A notable exception was the presence of detectable levels of CSF hsCRP in a subset of subjects with critical and severe COVID-19 illness, stroke and encephalopathy, which correlated with the magnitude of corresponding serum increase. The upsurge of CSF hsCRP in COVID-19 subjects suggests that such acute phase reactant is linked to the mechanisms of stroke and ischemia during acute and subacute stages of COVID-19. It is uncertain if CRP in CSF is actively or passively transported from serum, due to brain endothelial pathology or from brain disease processes, as neurons may have capability to produce such pentraxin. (55) Future studies should focus on determining the role of CRP in CSF, establishing the outcome of neurological disease in COVID-19 subjects when CSF hsCRP is detected as its presence may have long-term implications in mechanisms of neurodegeneration(55). Surprisingly, levels of CSF ferritin and D-dimer, showed no significant increase and/or did not mirror the marked elevation observed in the serum levels in COVID-19 subjects. CSF ferritin and D-dimer levels were elevated in COVID-19 cases mostly due to the presence of cases of SAH or stroke with ICH. Remarkably, standard assays for quantification of fibrinogen and factor VIII in CSF failed to detect such analytes in both COVID-19 and control cases including stroke cases, findings that suggest either the absence of such molecules in the CSF or the lack of sensitivity of the assay for their detection.

Although the prospect of a consensus for common CSF signatures in patients with neurological manifestations of COVID-19 is challenged by the diversity of clinical presentations, patient heterogeneity, overlapping risk factors and co-morbidities, our study has further implications for the understanding of the neuropathogenesis of these neurological complications. The paucity of neuroinflammatory changes in COVID-19 CSF rejects the prevailing assumption that pro-inflammatory cytokines, primary neuroinflammation or SARS-CoV2 neurovirulence are the main drivers of neuropathogenesis in COVID-19. We believe the few identified “neuroinflammatory” processes in the CSF of COVID-19 (23, 24, 56) are mostly derived from homeostatic neuroglial responses by microglia and astroglia to systemic pathology such as ischemia, hypoxia or systemic critical illnesses(57, 58) rather than active inflammation or neurovirulence. However, in critical COVID-19 and some encephalopathy cases there are increased CSF markers of neuroaxonal injury such as NF-L, supporting the view that CNS damage does occur in subsets of patients with COVID-19. The pathologic mechanisms underlying this CNS damage remain unknown, though may be secondary to microischemic injuries, severe systemic inflammation, and/or systemic metabolic dysfunction. Future postmortem and animal studies will be helpful to elucidate these mechanisms. Another interesting future direction will be to investigate the potential role of SARS-CoV2 antibodies in the pathogenesis of acute, subacute or even long-term consequences of COVID-19 and effect on CNS function including in those cases of the reported “brain fog” and other neurological symptoms in the so-called “long term haulers” (59).

The strengths of this study include a comprehensive analysis of markers of disease immunopathogenesis in the CSF from COVID-19 subjects as compared with CSF from healthy controls, stroke and neuroinflammatory disorders subjects, and a comprehensive clinical, neuroimaging and laboratory phenotyping of the subjects, but a few limitations are important to mention. First, we mainly evaluated cytokines and immune factors which were selected based on their previously reported relevance to COVID-19. This was a limited study of paired CSF-serum samples for cytokine and antibody profiling. This was a necessary impediment because the limited availability of CSF and blood samples for research purposes during the emergency situation as well as the financial constraints for expanding the scope of testing. Moreover, during the period of the study, blood sampling in many patients with COVID-19 in a critical or severe stage was restricted to standard diagnostic testing. Second, this study is limited to the clinical experience in a tertiary referral center, a relatively small cohort of patients accrued during a short period of time and a small sample size for the relatively high number of comparisons. Finally, and more generally, future multi-center studies should include well acquired and phenotyped CSF repository samples that allow the expansion of analytical approaches of the CSF to determine pathogenic factors involved in the neurological complications during acute and long-term consequences of COVID-19.

## METHODS

### Study design

The CSF characteristics of a cross-sectional cohort of 18 hospitalized adult COVID-19 patients with neurological manifestations, were compared to those of 14 age matched healthy and 68 non-COVID-19 neurological disease controls to investigate immune and neuroinflammatory changes in the CSF which may be associated with pathogenesis of neurological involvement. The sample size of the COVID-19 group was determined by convenience and it corresponds to all COVID-19 patients that underwent CSF analysis during the period of study at our institution. The COVID-19 CSF was collected from patients undergoing standard of care evaluation for COVID-19 neurological complications at a tertiary care academic center during the period April 1-July 31 2020. Only patients with complete record of neurological examination by a neurologist, neuroimaging and NS-NAAT and/or immunological diagnosis of COVID-19 were included. The CSF was examined for 1) the presence of SARS-CoV2 virus, 2) antibody responses against SARS-CoV2, 3) selected cytokines associated with the systemic inflammatory response in COVID-19,(35-38) 4) markers of coagulation and acute phase reactants, and 5) NF-L, a marker of neuroaxonal damage. COVID-19 diagnosis was based on a positive NS-NAAT or demonstration of serum anti-SARS-CoV2 IgG or IgA antibodies. Clinical information was gathered through standardized interviews with patients or patient surrogates, or through electronic medical record (EMR) review if no interview was possible. Complete neurological examination, neuroimaging and laboratory data were recorded in a REDCap database of the Johns Hopkins Division of Neuroimmunology and Neuroinfectious Diseases-CSF Biorepository (NINI-CSFBiorep).

### Clinical definitions for COVID-19 group

Neurological manifestations in COVID-19 were categorized in three diagnostic groups: stroke, encephalopathies and headaches/others. COVID-19 stroke cases included subjects with ischemic stroke from intracranial atherosclerosis, cardioembolic, small vessel disease and other causes (34) and/or hemorrhagic stroke including intracerebral and subarachnoid hemorrhages confirmed by clinical and neuroimaging assessment. COVID-19 encephalopathy diagnosis included subjects with diffuse neurological dysfunction with altered consciousness with change in cognition and/or with a perceptual disturbance not better accounted for by a pre-existing or evolving chronic dementia (60) or sedation without evidence of stroke. Subjects with headache without mental status changes, with or without cranial nerve involvement without evidence of stroke or other structural lesions were classified in the group of headaches/other. COVID-19 disease severity was based on National Institutes of Health (NIH) Guidelines (61) and defined as follows: 1) Critical illness: respiratory failure, septic shock, and/or multiple organ dysfunction 2) Severe illness: respiratory frequency >30 breaths per minute, oxygen saturation (SaO2) ≤93% on room air at sea level, a ratio of arterial partial pressure of oxygen to fraction of inspired oxygen (PaO_2_ /FiO_2_) <300 mmHg or lung infiltrates >50%; 3) Moderate illness: Evidence of lower respiratory disease by clinical assessment or imaging and SaO2 >93% on room air at sea level and 4) Mild Illness: Individuals who had any of various signs and symptoms (e.g., fever, cough, sore throat, malaise, headache, muscle pain) without shortness of breath, dyspnea, or abnormal imaging. We determined the presence of COVID-19 hyperinflammatory syndrome (C-HIS) based on the clinical profile and combination of markers of systemic inflammation (e.g., ferritin, D-dimer, CRP and IL-6)(27). To determine the effect of time to CSF sampling as related with period of infection, two groups were established: An “early” CSF collection group for samples obtained within 8 days of the first positive NS-NAAT, and a “late” CSF collection group for samples obtained 9 days or after(62).

### Clinical definitions for control groups

Convenience CSF control group samples collected prior to the COVID-19 pandemic were used. The CSF samples were derived from newly diagnosed, treatment naï ve and well characterized subjects available at the NINI-CSFBiorep and selected with the best attempt to match by age (+/-5 years) the COVID-19 CSF samples. CSF control samples were classified as: 1) healthy controls, comprised by subjects with normal neurological examination and normal brain MRI who underwent evaluation for headaches or pseudotumor cerebri; 2) acute infectious meningitis, 3) acute viral encephalitis (e.g., herpes simplex, varicella-zoster encephalitis)(30), 4) autoimmune encephalitis (31), 5) NMO (32), 6) neurosarcoidosis (33), and 7) stroke, which included subjects with ischemic stroke (34) preceding the COVID-19 period.

### Laboratory studies

#### SARS-CoV2 virus and anti-SARS-CoV2 antibody detection in CSF

NAAT of SARS-CoV2 RNA in CSF was performed by RT-PCR. Two regions of the nucleocapsid (N) gene (N1 and N2) were used as assay targets per the FDA Emergency Use Authorization package insert (https://www.fda.gov/media/134922/download). ddPCR was used to confirm the results on a subset of the specimens (https://www.fda.gov/media/137579/download). The human RNase P gene (RP) was the internal control for both assays(63). Quantification of anti-SARS-CoV2 IgG and IgA antibodies used a previously validated ELISA kit (Euroimmune, Germany)(64) which identify antibodies against subunit 1 of the trimeric SARS-CoV2 spike protein. The cutoff for positivity was 1.23 units for IgG and 5 units for IgA as established previously (64).

#### Cytokine profiling

Quantification of the cytokines IL6, TNFa, IFNγ, IL-10, IL12p70 and IL17a was performed using the Simoa™. Cytokine 6-plex panel array assay using a Quanterix HD-X ^®^ analyzer. CSF NFL was measured using the Simoa™ NF-Light Kit (Quanterix Corporation, Lexington, MA, USA) on the Quanterix HD-X^®^ platform.

#### Acute phase reactants and coagulation markers

Ferritin and hsCRP were measured on Roche Diagnostics Cobas c 701 and e 801 analyzers, respectively. Fibrinogen quantification used a clot-based assay (Siemens, Marburg Germany). D-dimer was measured by an immunoturbidimetric assay (Innovance D-Dimer, Siemens, Marburg, Germany). Factor VIII assessment used a chromogenic Assay (Chromogenix, Bedford, MA).

### Statistics

Continuous variables were described using medians and interquartile ranges, while categorical features with percentages. Planned comparisons between COVID-19 and control groups were performed using Mann-Whitney test. All 3 COVID-19 diagnostic categories were compared with each control group and with each other. For CSF hsCRP and D-dimer, outputs were dichotomized with 0.2 mg/L as the cut-off considering the bimodal concentrations found; An initial global test assessed differences in groups and was followed by individual Fisher’s exact tests for comparison among groups. For analysis of cytokines, values with a coefficient of variation higher than 30% were disregarded. Missing concentration values below the lower limit of detection were calculated by dividing the lower limit of quantification (LLOQ) corresponding for each cytokine by the square root of 2. Spearman’s correlation coefficient (Rho; ρ) was evaluated as well for relating NF-light concentrations with the other immunomarkers. Significant p values were set below 0.05. We specified our primary analyses as global tests comparing COVID-19 groups versus healthy and neurologic disease controls, and we considered our study to be exploratory in nature. As a result, we did not adjust for multiple comparisons. Analytes other than cytokines were analyzed with the obtained raw data. Statistical analysis was performed in Stata v.14. (StataCorp, Texas, USA).

### Study Approval

This study was approved by the Johns Hopkins Institutional Review Board (IRB) for longitudinal acquisition of clinical and biological samples in patients with neurological disorders. An informed consent was obtained from each patient or next-of-kin representative.

## Data Availability

De-identified data will be made available to qualified investigators upon written request to the corresponding author.

## AUTHOR CONTRIBUTIONS

CAP conceived, designed and supervised the overall study. CAP, MAG and PVB designed the overall study, acquired, organized and analyzed data. MAG, GP and KCF analyzed the data and performed statistical analysis. AM, AL, LS, TK, HM and MC conducted laboratory analysis, acquired and analyzed data. All other members of the Hopkins Neuro-COVID-19 Group contributed to the acquisition of clinical data by evaluating patients and acquiring biological samples. CAP, MAG and PVB drafted the manuscript, and all authors contributed to the discussion of results, revised and edited the manuscript.

## ACKNOWLEDGMENTS

We would like to thank all patients and/or their families for giving consent for clinical information and sample collection for this study. Additionally, we want to thank the clinical and laboratory teams for their assistance in the evaluation of patients and sample collection, as well as the hospital staff for their dedication in the fight of COVID-19. This work was supported by NIH R01-NS110122 and The Bart McLean Fund for Neuroimmunology Research. We gratefully acknowledge Drs. Lyle Ostrow and Eric Matthew Aldrich for their help with clinical evaluations of patients and Dr. Rebecca Gottesman for her advice about the study design and statistical analysis.

