## Supplemental Data for "Cerebrospinal fluid in COVID-19 neurological complications: no cytokine storm or neuroinflammation"

Maria A. Garcia et. al.

Supplemental Tables

Supplemental Figures

Study Group Membership

Supplemental Table 1.
**Demographic features and CSF characteristics of healthy and disease controls**

| **Group** | **Age** (median, IQR) years | **Sex** (male %) | **Cerebrospinal Fluid** | | | |
|---|---|---|---|---|---|---|
| | | | **WCC** >5 cells/µL (n) | **Protein** >45 mg/dL (n) | **hsCRP** >0.2 mg/L | **D-dimer** >0.19 mg/L |
| Healthy Controls N=14 | 67.5 (51–75) | 57.1% | 1/14 | 6/14 | 0/13 | 3/11 |
| Autoimmune encephalitis N=14 | 37.5 (23–53) | 42.8% | 5/14 | 3/14 | 1/13 | 1/8 |
| Acute meningitis N=12 | 50.5 (28-67.5) | 50% | 11/12 | 10/12 | 0/9 | 5/9 |
| Acute encephalitis N=11 | 62 (32–65) | 54.5% | 6/11 | 7/11 | 0/7 | 3/6 |
| Neuro-Sarcoidosis N=12 | 54 (47.5-65.5) | 33.4% | 7/12 | 9/12 | 0/10 | 7/9 |
| Neuro Myelitis Optica N=11 | 33 (26–48) | 19% | 6/11 | 5/11 | 0/7 | 2/7 |
| Stroke N=8 | 44 (39.5-50.5) | 75% | 3/8 | 5/8 | 0/8 | 4/7 |

Supplemental Table 2.
**Characteristics of the CSF as related with anti-SARS-CoV2 IgG and IgA antibodies**

| ID # | COVID-19 Diagnostic Group | CSF WCC cell/µL | CSF RBC cell/µL | CSF Protein mg/dL | IgG index | Q Alb | CSF Anti-SARS-Cov2 IgG Units[A] | CSF Anti-SARS-Cov2 IgA Units[B] |
|---|---|---|---|---|---|---|---|---|
| 1 | Stroke/SAH | 313 | 152000 | 56 | NA | NA | **3.68** | **10.26** |
| 2 | Stroke/ Ischemic | 0 | 15 | 5.5 | NA | NA | 0.76 | 0.42 |
| 3 | Stroke/SAH | 5 | 2000 | 85 | NA | NA | NA | NA |
| 5 | Stroke/ Ischemic/C- HIS | 18 | 6000 | 277 | NA | NA | **12.29** | 0.76 |
| 7 | Stroke/ Ischemic/C-HIS | 1 | 0 | 26 | 0.55[C] | 3.65 | **29.54** | **10.26** |
| 9 | Stroke/ Ischemic and ICH | 56 | 2000 | 63 | NA | NA | **5.11** | 0.47 |
| 4 | Encephalopathy/ C-HIS | 1 | 9 | 54 | NA | NA | **6.80** | 1.38 |
| 6 | Encephalopathy/ C-HIS | 25 | 1000 | 31 | NA | NA | **7.75** | **5.76** |
| 12 | Encephalopathy/ C-HIS | 0 | 0 | 52 | NA | NA | **1.36** | 0.62 |
| 13 | Encephalopathy/ C-HIS | 1 | 99 | 56 | 0.62[C] | 7.95 | 0.66 | 0.90 |
| 15 | Encephalopathy | 1 | 0 | 31 | 0.43[C] | 3.84 | 0.03 | 0.30 |
| 16 | Encephalopathy | 1 | 160 | 62 | 0.72[C] | 8.88 | **4.12** | 1.59 |
| 17 | Encephalopathy | 3 | 0 | 30 | 0.38 | 4.53 | **16.12** | 3.17 |
| 8 | Headache/other | 1 | 109 | 23 | NA | NA | **3.76** | 0.50 |
| 10 | Headache/ other | 3 | 0 | 33 | NA | NA | **1.60** | 0.13 |
| 11 | Headache/ other | 0 | 1 | 35 | NA | NA | **2.93** | **5.04** |
| 14 | Headache/ other | 3 | 0 | 27 | 0.52[C] | 5.06 | **6.92** | 1.46 |
| 18 | Headache/ other | 1 | 27 | 27 | 0.50 | 4.02 | 0.06 | 0.03 |

[A] Cutoff for IgG positivity at 1.23 units. Positive values are in bold.
[B] Cutoff for IgG positivity at 5.0 units. Positive values are in bold.
[C] Oligoclonal bands tested negative in CSF and serum
*Abbreviations:* SAH: subarachnoid hemorrhage; Q Alb: albumin quotient; ICH: intra-cerebral hemorrhage; C-HIS: COVID-19 hyper inflammatory syndrome

Supplemental Table 3.
**Percentage of detection of CSF biomarkers in CSF from COVID-19 and controls**

| **Analyte/Group** % analyte detection | **Healthy controls** (n=12 ) | **COVID-19** (n=18) | **Acute meningitis** (n=12 ) | **Autoimmune Encephalitis** (n=11) | **Acute Encephalitis** (n=11) | **NMO** (n=11) | **Neuro-Sarcoidosis** (n=11) | **Stroke** (n=8) |
|---|---|---|---|---|---|---|---|---|
| **IL-6** (LLQ=0.017 pg/mL) | 12<br>100.0% | 18<br>100.0% | 12<br>100.0% | 10<br>90.9% | 10<br>90.9% | 10<br>90.9% | 11<br>100.0% | 6<br>75.0% |
| **IL-10** (LLQ=0.03 pg/mL) | 10<br>83.3% | 15<br>83.3% | 10<br>83.3% | 9<br>81.8% | 6<br>54.5% | 6<br>54.5% | 11<br>100.0% | 4<br>50.0% |
| **TNFα** (LLQ= 0.018 pg/mL) | 11<br>91.7% | 17<br>94.4% | 12<br>100.0% | 10<br>90.9% | 9<br>81.8% | 9<br>81.8% | 11<br>100.0% | 6<br>75.0% |
| **IFNγ** (LLQ=0.082 pg/mL) | 11<br>91.7% | 16<br>88.9% | 11<br>91.7% | 10<br>90.9% | 10<br>90.9% | 11<br>100.0% | 11<br>100.0% | 7<br>87.5% |
| **IL12p70** (LLQ=0.02 pg/mL) | 7<br>58.3% | 13<br>72.2% | 11<br>91.7% | 8<br>72.7% | 6<br>54.5% | 7<br>63.6% | 10<br>90.9% | 5<br>62.5% |
| **IL-17A** (LLQ=0.003 pg /mL) | 11<br>91.7% | 18<br>100.0% | 12<br>100.0% | 9<br>81.8% | 9<br>81.8% | 10<br>90.9% | 11<br>100.0% | 8<br>100.0% |
| **NF-light** (LLQ= 1.6 pg/mL) | 12<br>100.0% | 18<br>100.0% | 12<br>100.0% | 10<br>90.9% | 11<br>100.0% | 11<br>100.0% | 11<br>100.0% | 8<br>100.0% |
| **Ferritin** ng/mL | 11<br>91.7% | 17<br>94.4% | 9<br>75% | 10<br>90.9% | 7<br>63.6% | 7<br>63.6% | 9<br>81.8% | 7<br>87.5% |
| **D-dimer** (LLQ= 0.19 mg/L | 10<br>83.3% | 15<br>83.3% | 9<br>75% | 6<br>54.5% | 6<br>54.5% | 7<br>63.6% | 8<br>72.7% | 8<br>100.0% |
| **hs-CRP** (LLQ=0.2 mg/L | 11<br>91.7% | 17<br>94.4% | 9<br>75% | 10<br>90.9% | 7<br>63.6% | 7<br>63.6% | 9<br>81.8% | 8<br>100.0% |

*Abbreviations:* LLQ: lower limit of quantification

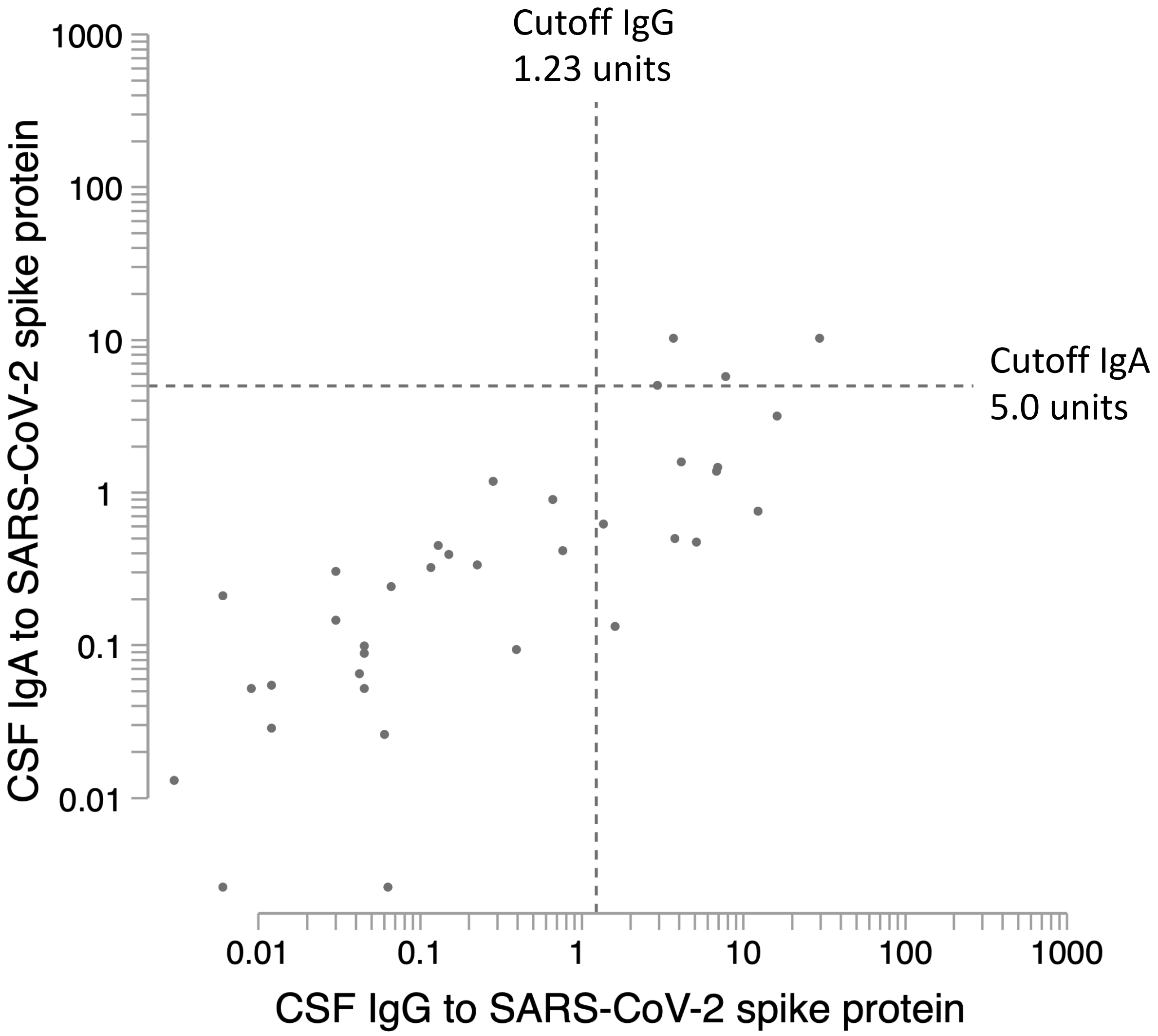


**Supplementary Figure 1.**

Scatter plot below distribution of the SARS-CoV2 IgG and IgA antibody measurements in the 37 CSF samples testes, 17 COVID-19 cases and 30 controls. Thirteen of 37 cases tested were positive, all of them COVID-19 cases. IgA was positive only in 4 of the 37 cases, and all of these 4 were also COVID-19 cases positive for IgG. The dotted line represents the cutoff for positivity: 1.23 units for IgG and 5 units for IgA. The scale is logarithmic.

Supplemental Table 4.
Johns Hopkins Neuro-COVID-19 Group

| Name | | Location | Contribution |
|---|---|---|---|
| Andrew Agostini | M.D. | Department of Neurology, Johns Hopkins University School of Medicine. | Evaluation of patients, acquired clinical data, and revised the manuscript for intellectual content |
| Mona Bahout | M.D, Ph.D. | Department of Neurology, Johns Hopkins University School of Medicine. | Evaluation of patients, acquired clinical data, and revised the manuscript for intellectual content |
| Srishti Bhagat | M.D. | Department of Neurology, Johns Hopkins University School of Medicine. | Evaluation of patients, acquired clinical data, and revised the manuscript for intellectual content |
| Pavan Bhargava | M.B.B.S., M.D. | Department of Neurology, Johns Hopkins University School of Medicine. | Evaluation of patients, acquired clinical data, and revised the manuscript for intellectual content |
| Sung-Min Cho | D.O., M.H.S. | Department of Neurology, Division of Cardiac Surgery, Cardiac Surgical Intensive Care, Johns Hopkins University School of Medicine. | Evaluation of patients, acquired clinical data, and revised the manuscript for intellectual content |
| Camilo Diaz Cruz | M.D. | Department of Neurology, Johns Hopkins University School of Medicine. | Evaluation of patients, acquired clinical data, and revised the manuscript for intellectual content |
| Sachin Gadani | M.D, Ph.D. | Department of Neurology, Johns Hopkins University School of Medicine. | Evaluation of patients, acquired clinical data, |
| Rebecca Gottesman | M.D. Ph.D. | Department of Neurology, Johns Hopkins University School of Medicine. | Provided advice about study design, statistical analysis and revised the manuscript for intellectual content |
| Brittney M.Howard | M.S. | Department of Pathology, | Laboratory testing, acquired laboratory data and revised the |

| | | | |
|---|---|---|---|
| | | Johns Hopkins University School of Medicine | manuscript for intellectual content |
| George Kannarkat | M.D., Ph.D. | Department of Neurology, Johns Hopkins University School of Medicine. | Evaluation of patients, acquired clinical data, and revised the manuscript for intellectual content |
| Amir Kheradmand | M.D. | Department of Neurology, Johns Hopkins University School of Medicine. | Evaluation of patients, acquired clinical data, and revised the manuscript for intellectual content |
| Michael Kornberg | M.D., M.S., Ph.D. | Department of Neurology, Johns Hopkins University School of Medicine. | Evaluation of patients, acquired clinical data, and revised the manuscript for intellectual content |
| Rafael Llinas | M.D | Department of Neurology, Johns Hopkins University School of Medicine. | Evaluation of patients, acquired clinical data, and revised the manuscript for intellectual content |
| Justin McArthur | M.B.B.S., M.P.H | Department of Neurology, Johns Hopkins University School of Medicine. | Evaluation of patients, acquired clinical data, and revised the manuscript for intellectual content |
| Bipasha Mukherjee | M.D. | Department of Neurology, Johns Hopkins University School of Medicine. | Evaluation of patients, acquired clinical data, and revised the manuscript for intellectual content |
| John C. Probasco | M.D | Department of Neurology, Johns Hopkins University School of Medicine. | Evaluation of patients, acquired clinical data, and revised the manuscript for intellectual content |
| Alexandra Simpson | M.D | Department of Neurology, Johns Hopkins University School of Medicine. | Evaluation of patients, acquired clinical data, and revised the manuscript for intellectual content |
| Jose Ignacio Suarez | M.D | Departments of Neurology and Neurosurgery, Anesthesiology and Critical Care Medicine, Johns Hopkins | Evaluation of patients, acquired clinical data, and revised the manuscript for intellectual content |

| | | | |
|---|---|---|---|
| | | University School of Medicine. | |
| Victor C. Urrutia | M.D. | Department of Neurology, Johns Hopkins University School of Medicine. | Evaluation of patients, acquired clinical data, and revised the manuscript for intellectual content |
| Arun Venkatesan | M.D. Ph.D. | Department of Neurology, Johns Hopkins University School of Medicine. | Evaluation of patients, acquired clinical data, and revised the manuscript for intellectual content |
| Wendy Ziai | M.D | Departments of Neurology and Neurosurgery, Anesthesiology and Critical Care Medicine, Johns Hopkins University School of Medicine. | Evaluation of patients, acquired clinical data, and revised the manuscript for intellectual content |
